# Clinical care and therapeutic practice of patients with neovascular age-related macular degeneration (nAMD) and diabetic macular edema (DME) and effectiveness of faricimab in treatment-naive patients in Germany - ZEUS research project

**DOI:** 10.1101/2025.03.26.25324686

**Authors:** Albert J Augustin, Finn Fabian Theine, Hakan Kaymak, Julia Clauß, Thilo Schimitzek, Max Zortel, Sandra Bluemich, Astrid Sader-Moritz

## Abstract

**Background:** Neovascular age-related macular degeneration (nAMD) and diabetic macular edema (DME) represent the leading causes of vision impairment and blindness among the elderly population and people with diabetes, respectively. To reduce disease burden and improve disease management, highly effective therapies and optimal treatment are of major importance. In September 2022, faricimab was approved for the treatment of nAMD and DME in Germany, allowing a treatment approach with dual Ang-2/VEGF-inhibition. This research project was conducted to investigate the clinical care and therapeutic practice in German nAMD and DME patients. Furthermore, by evaluating upload patterns in real-world practice, this study aimed to assess the use of faricimab in treatment-naive patients who received faricimab as a first-line treatment at these centers. Data on visual acuity and drying of the retina was collected in a descriptive manner.

**Methods:** ZEUS is a multicenter retrospective analysis of anonymized routinely collected data on patients with nAMD or DME who have been treated with intravitreal injections (IVT) between October 2022 and September 2024. A total of 24 sites (ophthalmologic practices or centers) across Germany reported predominant IVT regimen, drug upload strategies and diagnostic imaging techniques used in the routine care of patients with nAMD and DME in an electronic case report form (eCRF). For the subset of IVT injection-naive patients receiving faricimab data were extracted from patient charts and entered in an anonymized aggregated form into eCRF. Analysis included proportion of IVT injection-naive patients treated with faricimab in first-line, baseline characteristics of these patients/eyes, and reduction of fluid in the macula and changes in visual acuity during faricimab upload phase.

**Results:** The majority of sites (62.5%) applied a treat-and-extent (T&E) regimen, regardless of the chosen IVT. In case of faricimab the predominant upload scheme was 4 injections applied by 50% of sites, compared to three injections for other IVTs (58.3%) depending on the disease (nAMD/DME). OCT displayed the standard diagnostic at all sites (100.0%), other diagnostic procedures such as fluorescein angiography (FA, 62.5%) and OCT-A (25%) were applied less often. Out of the 24 sites, more than half initiated faricimab as first-line therapy in ≥20% of patients. More than 20% achieved a CRT reduction of either 40-60%, 20-40% or 0-20% during faricimab upload; a reduction of more than 80% was achieved in 14.4% of nAMD eyes. The majority of DME eyes (43.9%) displayed a CRT reduction of 20-40%. In the faricimab upload phase, absence of IRF was achieved in 67.2% of nAMD eyes, while an absence of only SRF and both IRF and SRF was seen in 65.0% and 56.7% of eyes. Approximately 80% of DME eyes were free of SRF in the upload phase. The main reason for using faricimab as first-line therapy was to achieve an increase in injection intervals and best effectiveness. Discontinuation rates were low.

**Conclusion:** This is the first analysis of general nAMD and DME treatment modalities as well as real-world effectiveness of faricimab in a large cohort of treatment-naive patients in Germany. Sites prefer individual approaches based on patients’ needs. First-line treatment with faricimab resulted in visual acuity gain during the faricimab upload phase in a real-world setting. These findings may help to better understand treatment strategies and first-line use of faricimab in nAMD and DME in Germany.

## INTRODUCTION

Neovascular age-related macular degeneration (nAMD) and diabetic macular edema (DME) represent leading causes of vision impairment and blindness among the elderly population and people with diabetes, respectively (Almony, 2023). Approximately 21 million people worldwide with about 350.000 patients in Germany suffer from DME or nAMD (Almony, 2023; G-BA, 2022b, 2022c; Li, 2020), with an expected increase in prevalence due to an aging population (Wong, 2014), which emphasizes the importance of treatment efficacy and optimal treatment to reduce disease burden and improve disease management.

nAMD is typically associated with a more rapid progression of vision loss as compared to the dry form (Stahl, 2020). If left untreated, patients with nAMD experience an average loss of three lines (15 letters) of visual acuity (VA) on the ETDRS scale in two years (Rosenfeld, 2006). Pathologically, choroidal neovascularization results in swelling, bleeding, and/or fibrosis (Ambati, 2012; Little, 2018), causing irreversible destruction of the macula and loss of the (sharp) central vision (fine detail vision) (Holekamp, 2019). DME is characterized by an accumulation of fluid in the extracellular space of the retina, particularly in the macular area, leading to blurred vision, distorted images, and central vision loss as a consequence of systemic vascular damage associated with underlying diabetes mellitus disease status (Zhang, 2022). Like nAMD, features of DME include increased vascular permeability, inflammation, intra- and epiretinal neovascularization, and an increased central retinal thickness (Almony, 2023).

The vascular endothelial growth factor (VEGF) stimulates angiogenesis and neovascularization, and increases vascular permeability (Rosenfeld, 2006). The introduction of therapies targeting vascular endothelial growth factor A (VEGF-A) has significantly impacted the clinical management of nAMD and DME by improving both, visual and anatomical outcomes (Brown, 2015; Chong, 2016; Khachigian, 2023; Nguyen, 2012; Virgili, 2017). Therefore, clinical guidelines for both, nAMD and DME include the recommendation of the use of intravitreal anti-VEGF injections (Bakri, 2019; Brown, 2017). However, due to the short duration of action of anti-VEGF agents and persistent disease activity, treatment usually requires frequent administration over several years to maintain therapeutic effect (Quah, 2024), placing a high treatment burden on patients and their caregivers (Wang, 2022).

Like VEGF, angiopoietin-2 (Ang-2) also plays a crucial role in neovascularization, vascular permeability, and additionally in inflammatory processes (Joussen, 2021). In human eyes, higher levels of Ang-2 correlate with disease severity in nAMD, reduced best-corrected VA and greater central macular thickness (Ng, 2017). Targeting multiple signaling pathways simultaneously allows to address multifactorial pathological processes as present in retinal diseases. The bispecific antibody faricimab (Roche/Genentech, Switzerland) was developed to inhibit both, Ang-2 and VEGF-A, and associated pathological processes. Phase III trials in patients with nAMD or DME demonstrated non-inferiority of faricimab to the anti-VEGF agent aflibercept with vision gains and anatomical improvements. This was achieved with faricimab injection intervals of up to 16 weeks (Heier, 2022; Quah, 2024). In September 2022 and August 2024, faricimab was approved for the treatment of adult nAMD/DME and ME due to RVO in Germany, respectively, allowing a treatment approach with dual Ang-2/VEGF-inhibition.

As results from clinical trials may not always translate to real-world clinical practice (Ciulla, 2021; Holz, 2015), studies evaluating treatment efficacy and safety of faricimab in a real world setting are of major importance. Existing real-world studies in nAMD (Kataoka, 2024; Khanani, 2023; Leung, 2023; Rush, 2023b) or DME (Kusuhara, 2023; Ohara, 2023; Rush, 2023a) primarily included patients with a prior treatment history. To our knowledge there is currently no data on the clinical care of treatment-naive patients treated with faricimab as first-line at centers and also practices across Germany.

This retrospective data collection aimed to evaluate the IVT treatment landscape of nAMD and DME in Germany, examining treatment regimens, imaging modalities, and clinical decision-making in ophthalmology practices. Additionally, this study was conducted to evaluate real-world utilization of faricimab in treatment-naive patients especially by evaluating upload patterns; data on visual acuity and drying of affected retina was collected in a descriptive manner.

## METHODS

### Study Design and Patients

ZEUS (DRKS00035007) is a multicenter retrospective analysis of anonymized routinely collected data on patients with nAMD or DME who have been treated with intravitreal injection therapies between October 2022 and September 2024.

The research project was conducted in full conformance with the Guidelines for good pharmacoepidemiology practice published by the International Society of Pharmacoepidemiology and local laws and regulations as applicable. The protocol and relevant supporting information were reviewed and approved by the primary voting ethical committee (i.e., Ethics committee of the Landesärztekammer Baden-Württemberg, F-2024-076) and submitted to local responsible committees, respectively.

A total of 24 ophthalmologic practices or centers across Germany provided the data pool for this aggregated analysis. No minimum follow-up time was required to participate in this study. Patients were included based on documented treatment initiation with faricimab, with a focus on those who had not received prior IVT therapy. This selection aimed to provide insights into first-line treatment effectiveness and reduce confounding factors related to previous anti-VEGF exposure.

To evaluate the predominant regimen for intravitreal therapy (IVT), drug upload strategies and diagnostic imaging techniques used in the routine care of patients with nAMD and DME, according site-specific questions were answered in the electronic case report form (eCRF). For the subset of IVT injection-naive patients receiving faricimab selected for the study by the investigator, data were extracted from patient charts and entered in an anonymized aggregated form into eCRF. At sites where several intravitreal injection-naive patients treated with faricimab in first line were eligible, patients with the longest observation period after faricimab injection were to be selected for inclusion in the research project. Investigators were allowed to include a maximum of 35 patients. Additional selection criteria were applied for IVT-naive patients receiving faricimab and are summarized in Table 1.

**Table 1:**
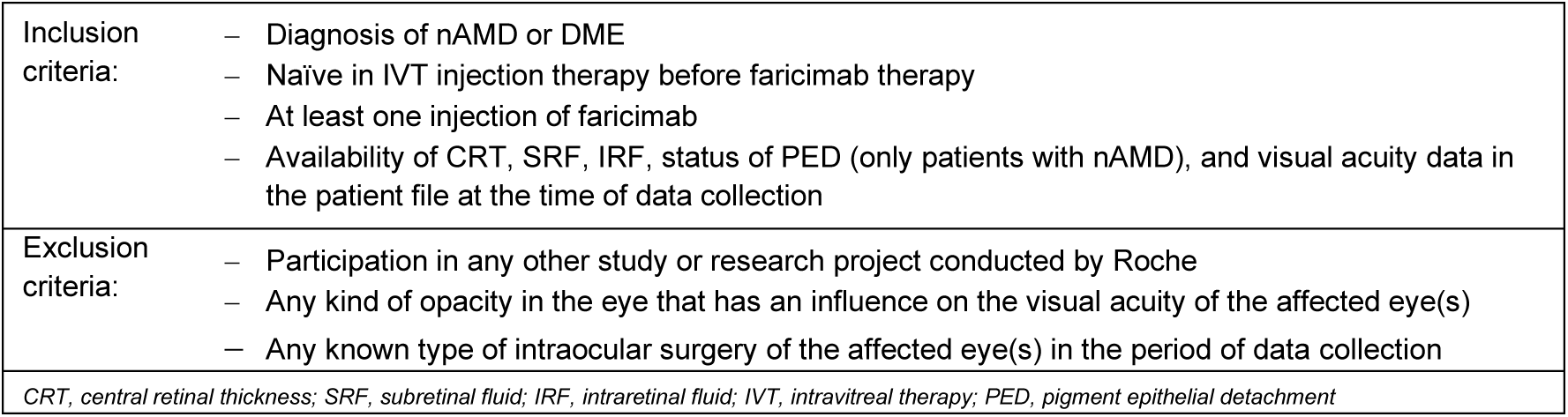
Inclusion and exclusion criteria.

|  |  |
| --- | --- |
| Inclusion criteria: | – Diagnosis of nAMD or DME<br>– Naïve in IVT injection therapy before faricimab therapy<br>– At least one injection of faricimab<br>– Availability of CRT, SRF, IRF, status of PED (only patients with nAMD), and visual acuity data in the patient file at the time of data collection |
| Exclusion criteria: | – Participation in any other study or research project conducted by Roche<br>– Any kind of opacity in the eye that has an influence on the visual acuity of the affected eye(s)<br>– Any known type of intraocular surgery of the affected eye(s) in the period of data collection |
| CRT, central retinal thickness; SRF, subretinal fluid; IRF, intraretinal fluid; IVT, intravitreal therapy; PED, pigment epithelial detachment |  |

### Objectives and endpoints

The primary objective was to evaluate the proportion of IVT injection-naive patients treated with faricimab in first-line in clinical practice of nAMD and DME from all faricimab treated patients. Secondary outcomes included patient and baseline characteristics of the affected eyes in IVT injection-naive patients (nAMD and DME) receiving first-line treatment with faricimab, reduction of fluid in the macula (assessed by central retinal thickness [CRT], subretinal fluid [SRF] and/or intraretinal fluid [IRF]), faricimab upload patterns (one injection could be documented as upload), the status of pigment epithelial detachment (PED; nAMD only) and VA (assessed by decimal best-documented VA [BDVA]) in affected eyes after faricimab treatment. Furthermore, the reason for faricimab therapy, treatment regimens, diagnostic imaging techniques, and therapy protocols used in clinical practice for nAMD and DME were evaluated.

### Statistical methods

No individual patient-level data were available for analysis, as patient data were collected in aggregated form. All variables, including originally continuous measures, were categorized to ensure consistency in reporting proportions as endpoints. Descriptive analyses were conducted to summarize the data. Categorical variables were presented as absolute frequencies and percentages. Missing values were not included in the calculation of percentages.

The site analysis set included all practices and centers that participated in the research project. For faricimab specific questions, all treatment-naive patients and eyes with nAMD and DME who were treated with faricimab and selected for inclusion by the investigator were included (patient and eye analysis set, respectively). Longitudinal analyses comprised only patients from the patient analysis set with completed faricimab upload as per physician’s definition. Baseline was defined as the date of the last assessment performed either at or before the first faricimab injection. All analyses of treatment-naive patients receiving first-line treatment with faricimab were conducted separately for the two populations of nAMD and DME patients, if not otherwise indicated.

No formal sample size calculation linked to hypothesis testing was conducted. No imputation methods were applied for missing data. The statistical evaluation was performed using the software package SAS release 9.4 (SAS Institute Inc., Cary, NC, USA).

## RESULTS

### Clinical care and therapeutic practice in German patients

Data on clinical care and therapeutic practice were available from all 24 participating sites. The majority of sites (62.5%) applied a treat-and-extent (T&E) regimen, regardless of the chosen IVT (Figure 1). Stratified by regimen, T&E was applied at 62.5%, 65.2%, 63.2%, 62.5% and 47.4% of sites for the treatment with aflibercept, ranibizumab, brolucizumab, faricimab and other IVT, respectively (Table 2). About one third of sites preferred a pro re nata (PRN) regimen, regardless of therapy. Accordingly, most sites (83.3%) stated that they did not apply different treatment regimens based on a specific IVT. With respect to uploading aflibercept, ranibizumab, brolucizumab and other IVTs, most sites (58.3%) did not distinguish between therapies and used three IVT injections (66.7%, 73.9%, 78.9%, 63.2%, respectively) (Figure 1; Table 3). In case of faricimab the predominant upload scheme was 4 injections applied by 50% of sites, followed by three (29.2%) and only one IVT injection (16.7%).

**Figure 1:**
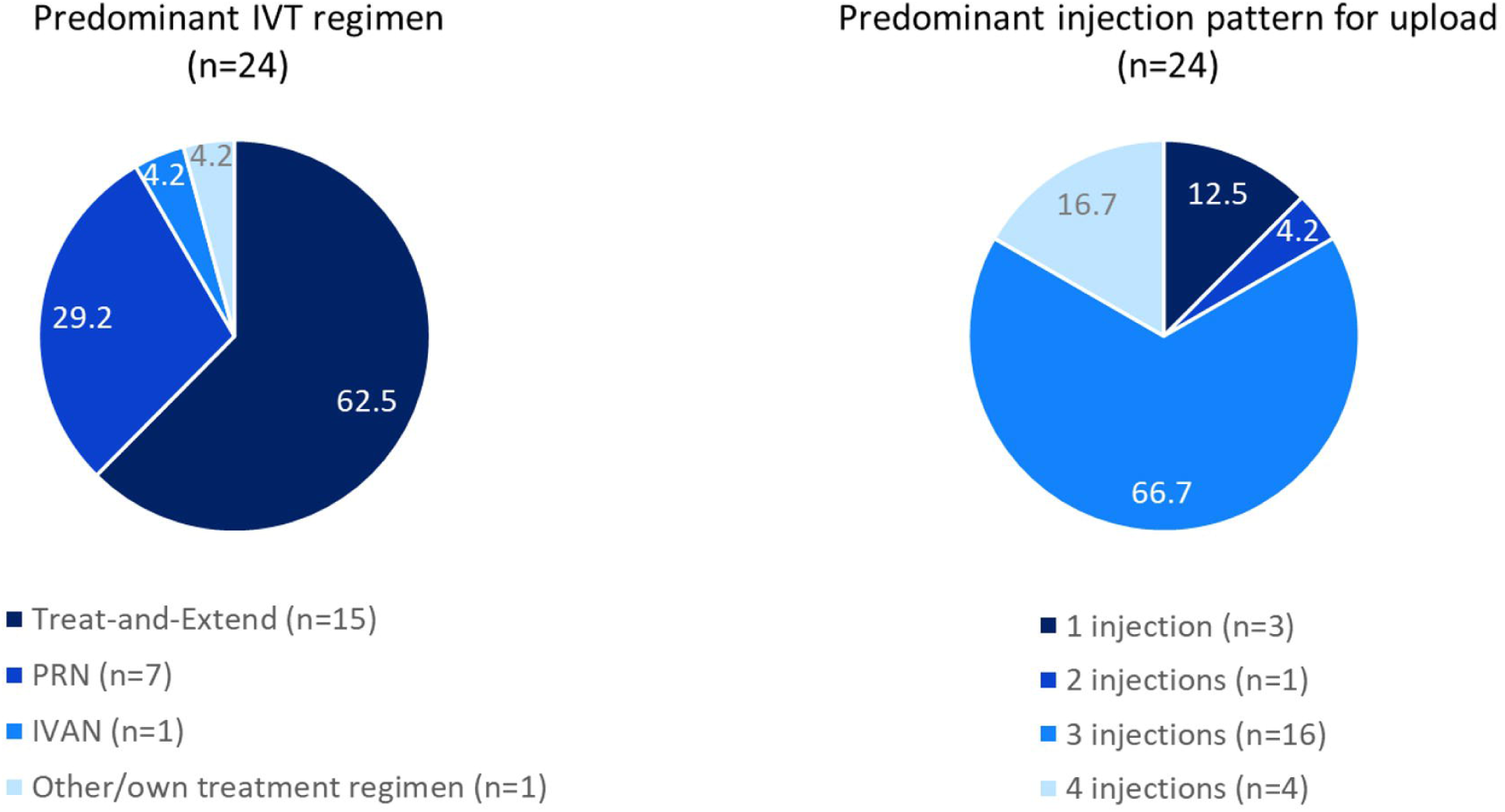
Predominant treatment regimens and upload patterns used, regardless of therapy. **N** refers to the number of sites. IVT: intravitreal therapy; **PRN:** pro re nata

**Table 2:**
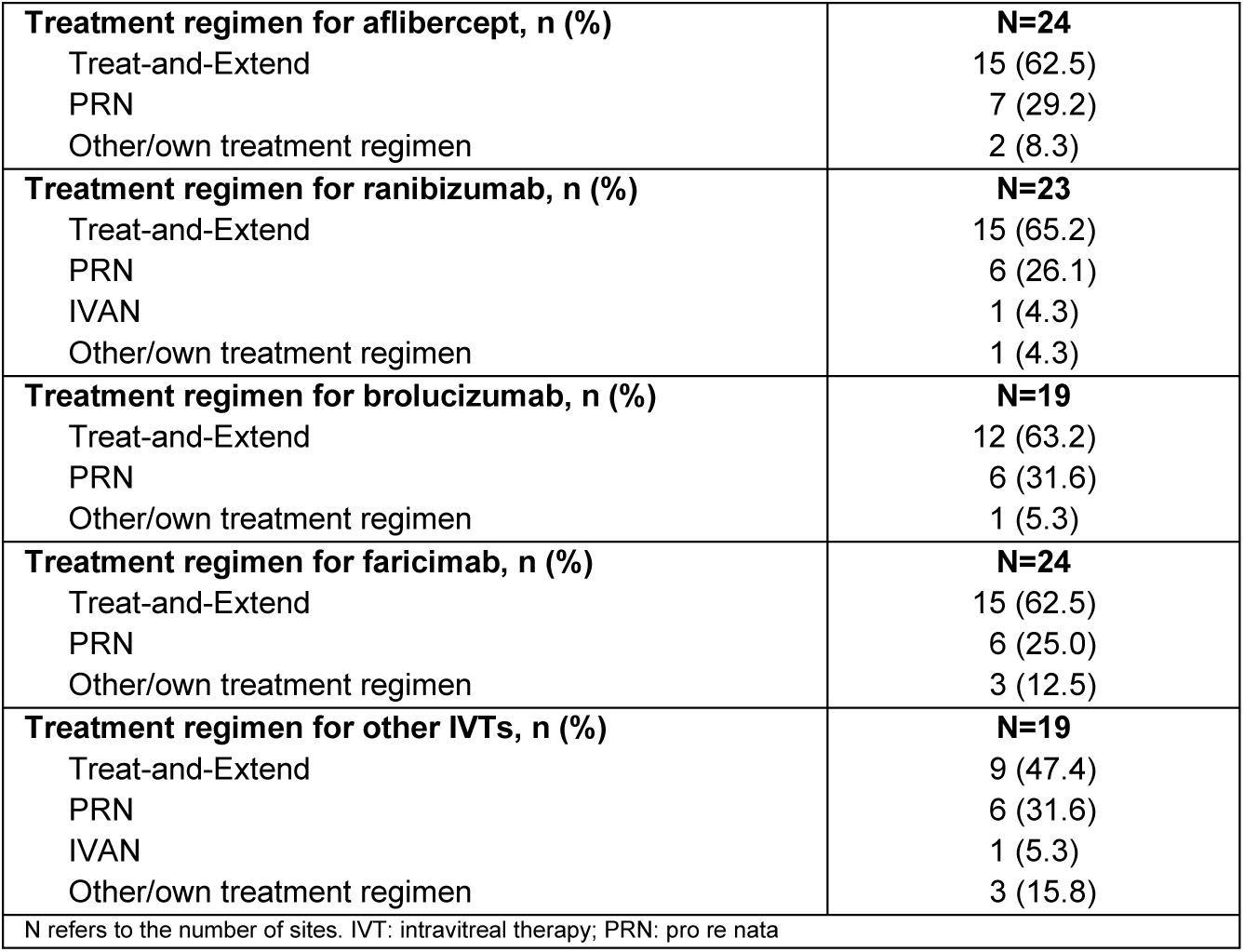
Treatment regimen used, based on therapy.

**Table 3:** Number of IVT injections based on therapy used.

|  |  |
| --- | --- |
| **IVT injection pattern for upload with aflibercept, n (%)** | **N=24** |
| 1 injection | 3 (12.5) |
| 2 injections | 1 (4.2) |
| 3 injections | 16 (66.7) |
| 4 injections | 4 (16.7) |
| **IVT injection pattern for upload with ranibizumab, n (%)** | **N=23** |
| 1 injection | 1 (4.3) |
| 2 injections | 1 (4.3) |
| 3 injections | 17 (73.9) |
| 4 injections | 4 (17.4) |
| **IVT injection pattern for upload with brolucizumab, n (%)** | **N=19** |
| 1 injection | 1 (5.3) |
| 2 injections | 1 (5.3) |
| 3 injections | 15 (78.9) |
| 4 injections | 2 (10.5) |
| **IVT injection pattern for upload with faricimab, n (%)** | **N=24** |
| 1 injection | 4 (16.7) |
| 2 injections | 1 (4.2) |
| 3 injections | 7 (29.2) |
| 4 injections | 12 (50.0) |
| **IVT injection pattern for upload with other IVTs, n (%)** | **N=19** |
| 1 injection | 3 (15.8) |
| 2 injections | 1 (5.3) |
| 3 injections | 12 (63.2) |
| 4 injections | 3 (15.8) |
| N refers to the number of sites. IVT: intravitreal therapy |  |

Regarding the imaging techniques employed at treatment initiation, OCT was the standard procedure at all sites (Figure 2). Other frequently used procedures included fluorescein angiography (FA) (62.5%), while OCT-A was applied less often (25%). After start of the therapy, first imaging OCT was most commonly performed after completion of the upload phase (58.3%). A smaller proportion of sites (20.8 %) conducted the first OCT imaging immediately after the first IVT injection, while 20.8 % of sites performed the imaging individually during the treatment course.

**Figure 2:**
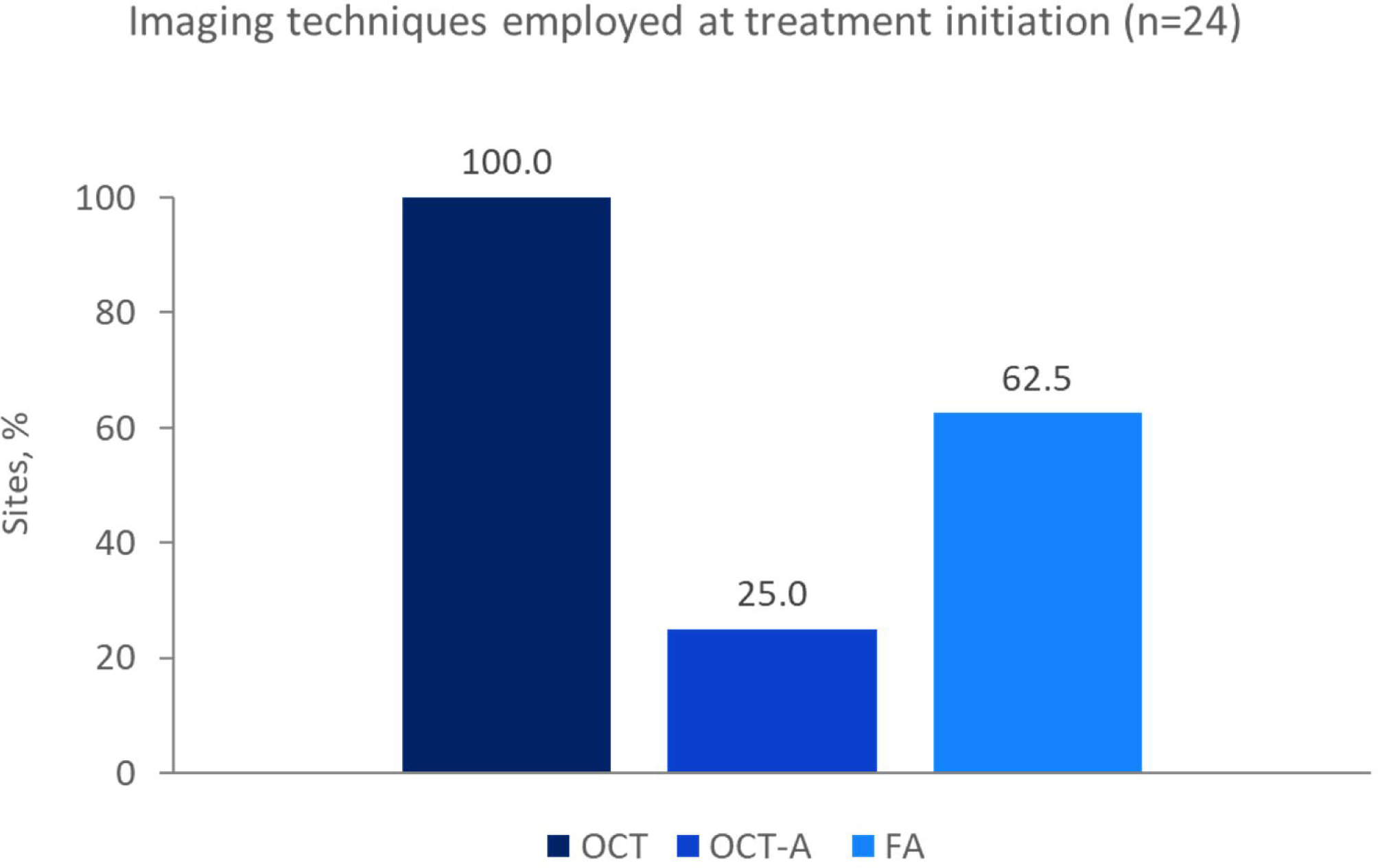
Imaging techniques employed at treatment initiation. N refers to the number of sites. FA, fluorescein angiography; OCT, optical coherence tomography; OCT-A, optical coherence tomography angiography

### Faricimab first-line treatment - Clinical characteristics of treatment-naive patients

A total of 3592 patients and 4287 eyes (nAMD and DME) were treated with faricimab in the participating clinical practices and centers. A subset of 228 treatment-naive patients (nAMD, 189 patients; DME, 39 patients) and 253 eyes (nAMD, 205 eyes; DME, 48 eyes) were included in the data analysis. Baseline patient demographics and eye characteristics are summarized in Table 4.

**Table 4:** Demographics and disease characteristics at baseline of our cohort of 228 individuals.

|  | nAMD | DME |
| --- | --- | --- |
| Number of treatment-naïve patients, n | 189 | 39 |
| **Gender, n (%)** |  |  |
| Male | 83 (43.9) | 28 (71.8) |
| Female | 106 (56.1) | 11 (28.2) |
| **Age group, n (%)** |  |  |
| <50 years | 1 (0.5) | 5 (12.8) |
| 50–60 years | 2 (1.1) | 8 (20.5) |
| 61–70 years | 26 (13.8) | 19 (48.7) |
| 71–80 years | 71 (37.6) | 3 (7.7) |
| >80 years | 89 (47.1) | 4 (10.3) |
| **Classification of CNV, n (%)** |  |  |
| Patients with occult CNV | 74 (39.2) | – |
| Patients with classic CNV | 60 (31.7) |  |
| Patients with RAP | 4 (2.1) |  |
| Patients with unknown classification | 54 (28.6) |  |
| **Type of diabetes, n (%)** |  |  |
| Diabetes type I | – | 10 (25.6) |
| Diabetes type II |  | 29 (74.4) |
| N refers to the number of patients. CNV: choroidal neovascularization; DME: diabetic macular edema; nAMD: neovascular age-related macular degeneration; RAP: retinal angiomatous proliferation |  |  |

Out of 24 sites, more than half initiated faricimab as first-line therapy in ≥20% of patients and about 30% in 10-19% (Figure 3). Considering only the treatment of nAMD patients, the majority of centers (more than 80%) applied faricimab in a first-line setting in more than 10% of patients. For DME patients, most centers (36.8%) applied faricimab as first-line in ≥20% of patients, while one in 5 centers used faricimab as a first-line treatment in less than 1% of patients.

**Figure 3:**
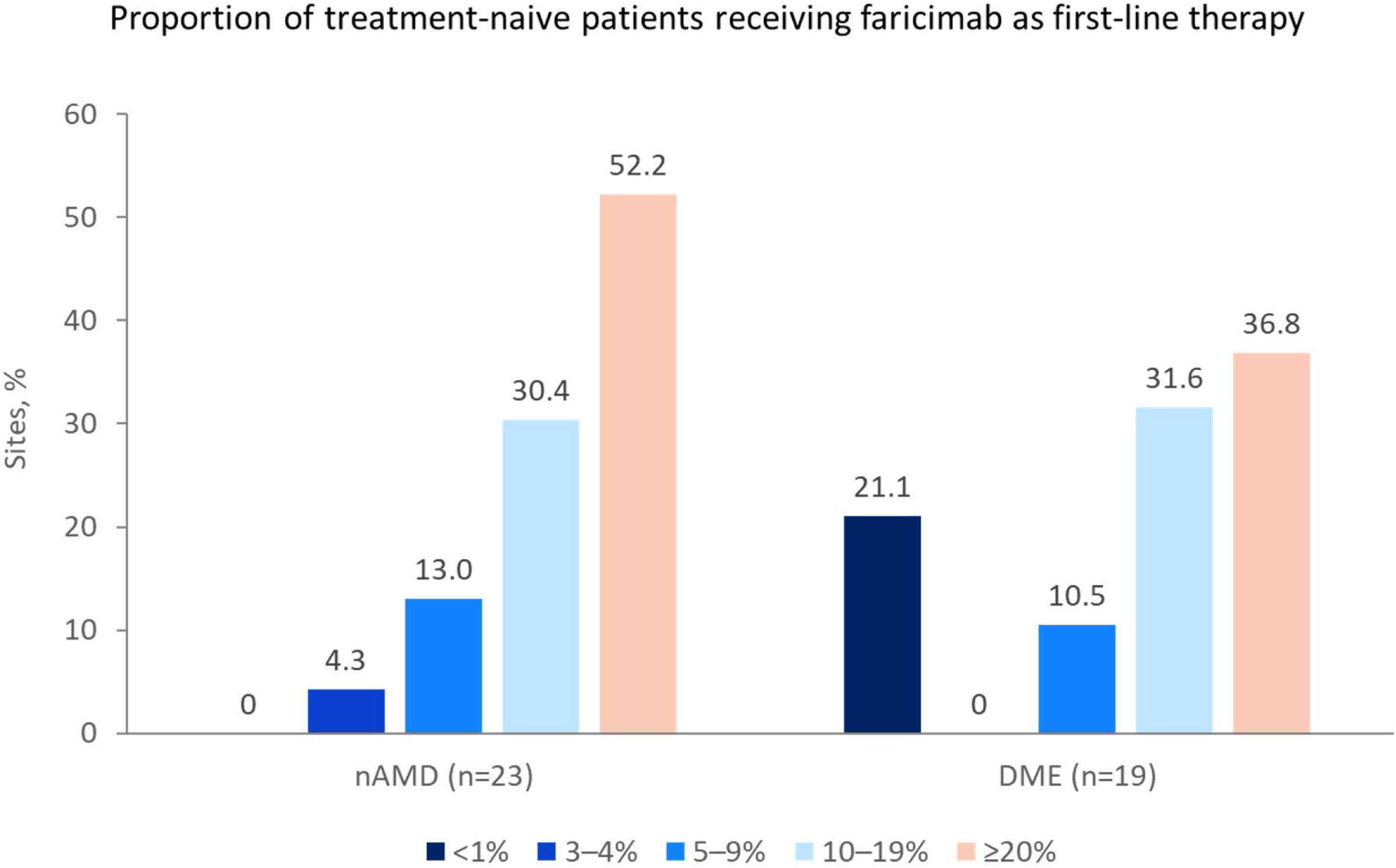
Proportion of treatment-naive patients receiving faricimab as first-line therapy. N refers to the number of sites. DME: diabetic macular edema; nAMD: neovascular age­ related macular degeneration

The subgroup of treatment-naive nAMD patients receiving faricimab as fist-line therapy included 56.1% females (Table 4). Most patients (47.1%) were older than 80 or aged between 71-80 years (37.6%), while few patients were younger than 61 years (1.6%). The proportion of nAMD patient with occult choroidal neovascularization (CNV) was 39.2%, while 31.7% of patients had classic CNV. 2% of nAMD patients suffered from retinal angiomatous proliferation (RAP). For approximately one in four patients (28.6%) CNV was not classified (Table 4). In the nAMD population, both intraretinal and subretinal fluid (IRF and SRF) was present in 39.5% of treatment-naive eyes. IRF only was reported in 28.3%, SRF only in 25.9% of eyes. Foveal and extrafoveal edema was present in 67.8% and 21.0% of nAMD eyes, respectively. Baseline BDVA was broadly distributed, particularly in the nAMD population, ranging from minor vision impairment to BDVA measures of 0.124-0.1 in 12.7% of patients (Table 5).

**Table 5:** Baseline characteristics of eyes.

|  | nAMD<br>(n=205) | DME<br>(n=48) |
| --- | --- | --- |
| **Pathology, n (%)** |  |  |
| Eyes with DRIL |  | 22 (45.8) |
| Eyes with rupture of ELM |  | 2 (4.2) |
| Eyes with unknown pathology |  | 3 (6.3) |
| **DME status, n (%)** |  |  |
| Patients with non-proliferative DME |  | 37 (77.1) |
| Patients with proliferative DME |  | 6 (12.5) |
| Patients with unknown DME status |  | 5 (10.4) |
| **Fluid location, n (%)** |  |  |
| Eyes with IRF | 58 (28.3) | 37 (77.1) |
| Eyes with SRF | 53 (25.9) | 4 (8.3) |
| Eyes with IRF and SRF | 81 (39.5) | 6 (12.5) |
| Eyes with foveal edema | 139 (67.8) | 46 (95.8) |
| Eyes with extrafoveal edema | 43 (21.0) | 7 (14.6) |
| PED | 88 (42.9) | – |
| **Visual acuity (decimal BDVA), n (%)** |  |  |
| 1 | 2 (1.0) | 3 (6.3) |
| 0.99–0.8 | 22 (10.7) | 3 (6.3) |
| 0.799–0.63 | 17 (8.3) | 8 (16.7) |
| 0.629–0.5 | 36 (17.6) | 10 (20.8) |
| 0.499–0.4 | 38 (18.5) | 10 (20.8) |
| 0.399–0.32 | 14 (6.8) | 1 (2.1) |
| 0.319–0.25 | 25 (12.2) | 5 (10.4) |
| 0.249–0.2 | 11 (5.4) | 2 (4.2) |
| 0.199–0.16 | 3 (1.5) | 0 (0.0) |
| 0.159–0.125 | 3 (1.5) | 0 (0.0) |
| 0.124–0.1 | 26 (12.7) | 4 (8.3) |
| 0.062–0.051 | 5 (2.4) | 0 (0.0) |
| <0.051 | 3 (1.5) | 2 (4.2) |
| N refers to the number of eyes. BDVA: best-documented visual acuity; DME: diabetic macular edema; DRIL: disorganization of retinal inner layers; ELM: external limiting membrane; IRF: intraretinal fluid; nAMD: neovascular age-related macular degeneration; PED: pigment epithelial detachment; SRF: subretinal fluid |  |  |

In the DME subgroup, patients were mostly male (71.8%). Nearly half of patients were aged between 61–70 years, followed by 20.5% in the age group of 50–60 years. Diabetes type II was more frequent than type I (74.4% vs 25.6%) (Table 4). Before start of faricimab therapy, a disorganization of retinal inner layers (DRIL) was present in 45.8% of eyes, while discontinuation of external limiting membrane (ELM) was displayed in 4.2% of eyes (Table 5). The retinal status of the majority of eyes (77.1%) was categorized as non-proliferative. In the DME population, the majority of eyes had IRF only (77.1%). Approximately one in ten eyes displayed SRF or both IRF and SRF. Foveal edema was observed in almost all eyes with DME (95.8%). The point of greatest fluid volume was CRT in 38.5% of nAMD and 41.7% of DME eyes. The majority of DME patients (>80%) displayed a decimal BDVA of 0.319-0.25 or better (Table 5).

The number of IVT injections used for upload in therapy-naïve faricimab patients is shown in Table 6. Most treatment-naive nAMD eyes (35.6%) received an upload of four faricimab injections. One or three faricimab injections were applied in 22.9% or 23.9% of eyes. Also in eyes with DME, upload mostly comprised four IVT injection. Five injections were applied in 10.4% of DME eyes. At the time of data cut-off, faricimab upload was not completed in 12.2% and 14.6% of nAMD and DME eyes, respectively. Thus, the analysis set including patients with completed faricimab upload as per physician’s definition comprised 221 eyes (nAMD: 180 eyes, DME: 41 eyes), and was used for longitudinal analyses.

**Table 6:** Number of IVT injections used for upload in therapy-naïve faricimab patients.

|  | **nAMD**<br>(n=205) | **DME**<br>(n=48) |
| --- | --- | --- |
| **Number of IVT injections used for upload, n (%)** |  |  |
| 1 injection | 47 (22.9) | 8 (16.7) |
| 2 injections | 7 (3.4) | 1 (2.1) |
| 3 injections | 49 (23.9) | 5 (10.4) |
| 4 injections | 73 (35.6) | 22 (45.8) |
| 5 injections | 4 (2.0) | 5 (10.4) |
| Upload not completed | 25 (12.2) | 7 (14.6) |
| N refers to the number of patients. DME: diabetic macular edema; nAMD: neovascular age-related macular degeneration |  |  |

### Effectiveness of faricimab first-line treatment in clinical practice

To assess the effectiveness of faricimab first-line treatment in nAMD and DME patients the following parameters were analyzed: the proportion of treatment-naive eyes with CRT reduction in pre-defined categories, absence of IRF and/or SRF and achieved changes in BDVA during the faricimab upload phase. Patients received a median of 3 (nAMD) or 4 (DME) upload injections, corresponding to a median observation period of approximately 12 weeks (nAMD), 16 weeks (DME).

In eyes with nAMD, more than 20% of eyes achieved a CRT reduction of either 40-60%, 20-40% or 0-20% during the faricimab upload phase (Figure 4). A reduction of more than 80% was achieved in 14.4% of nAMD eyes. The majority of eyes with DME (43.9%) displayed a CRT reduction of 20-40%. Compared to nAMD, fewer eyes achieved reductions of more than 60%. IRF was absent in 67.2% of nAMD eyes, while an absence of only SRF and both IRF and SRF was achieved in 65.0% and 56.7% of eyes (Figure 5). PED was absent in 60.0% of nAMD eyes. Approximately 80% of DME eyes were free of SRF. The evaluated eyes had a median improvement in visual acuity of 0.2-0.3 (decimal; nAMD and DME). Most nAMD eyes (34 %) displayed a change in decimal BDVA of ≤0.1, followed by 15.0-18.3% of eyes with changes in BDVA of up to 0.3-0.4. An improvement of >0.6 was seen in 7.2% of eyes during the upload phase. BDVA changes in DME eyes also ranged from ≤0.1 to >0.6 (Table 7).

**Figure 4:**
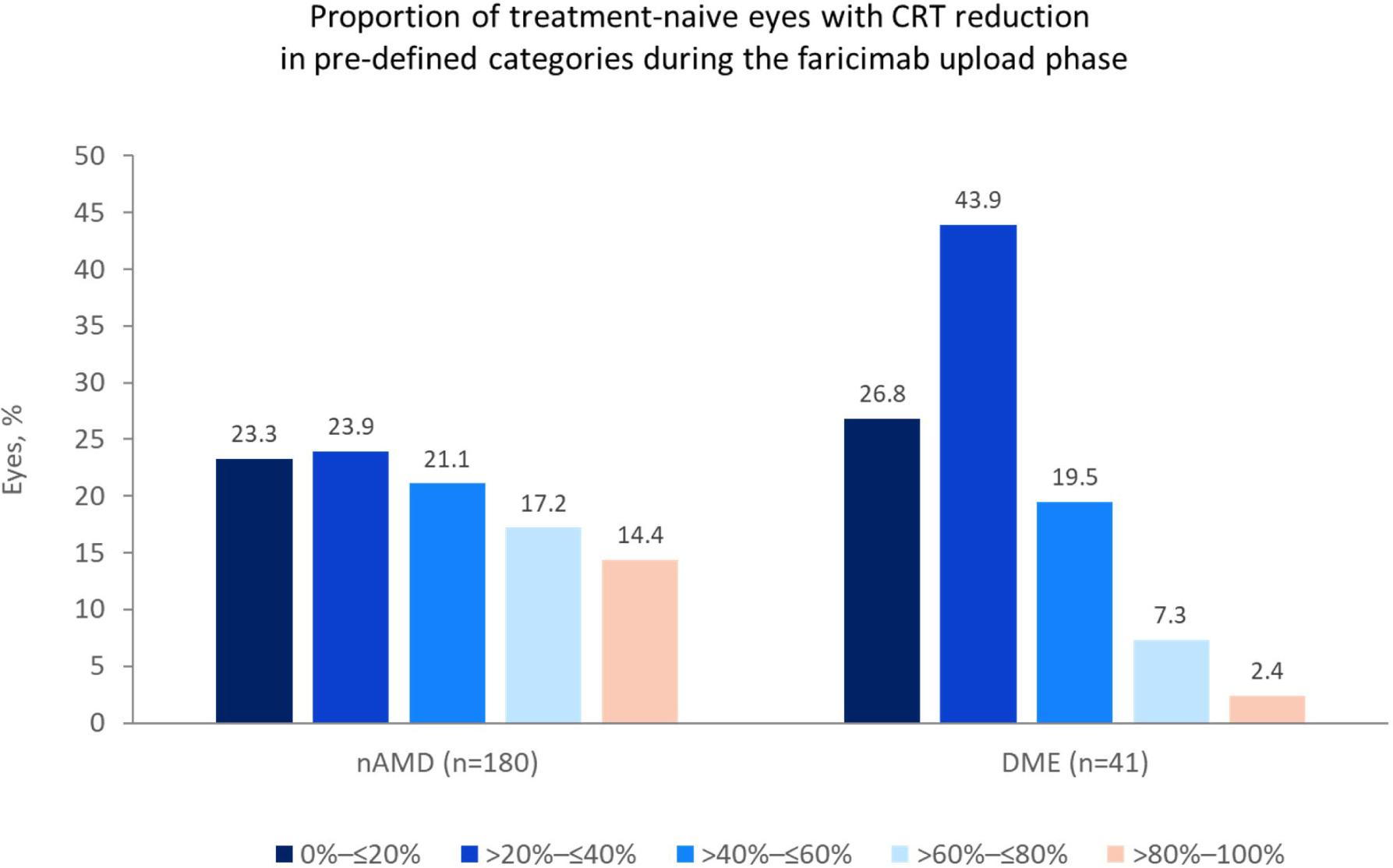
Proportion of treatment-naive eyes with CRT reduction in pre-defined categories during faricimab upload phase. N refers to the number of eyes. CRT: central retinal thickness; DME: diabetic macular edema; nAMD: neovascular age-related macular degeneration

**Figure 5:**
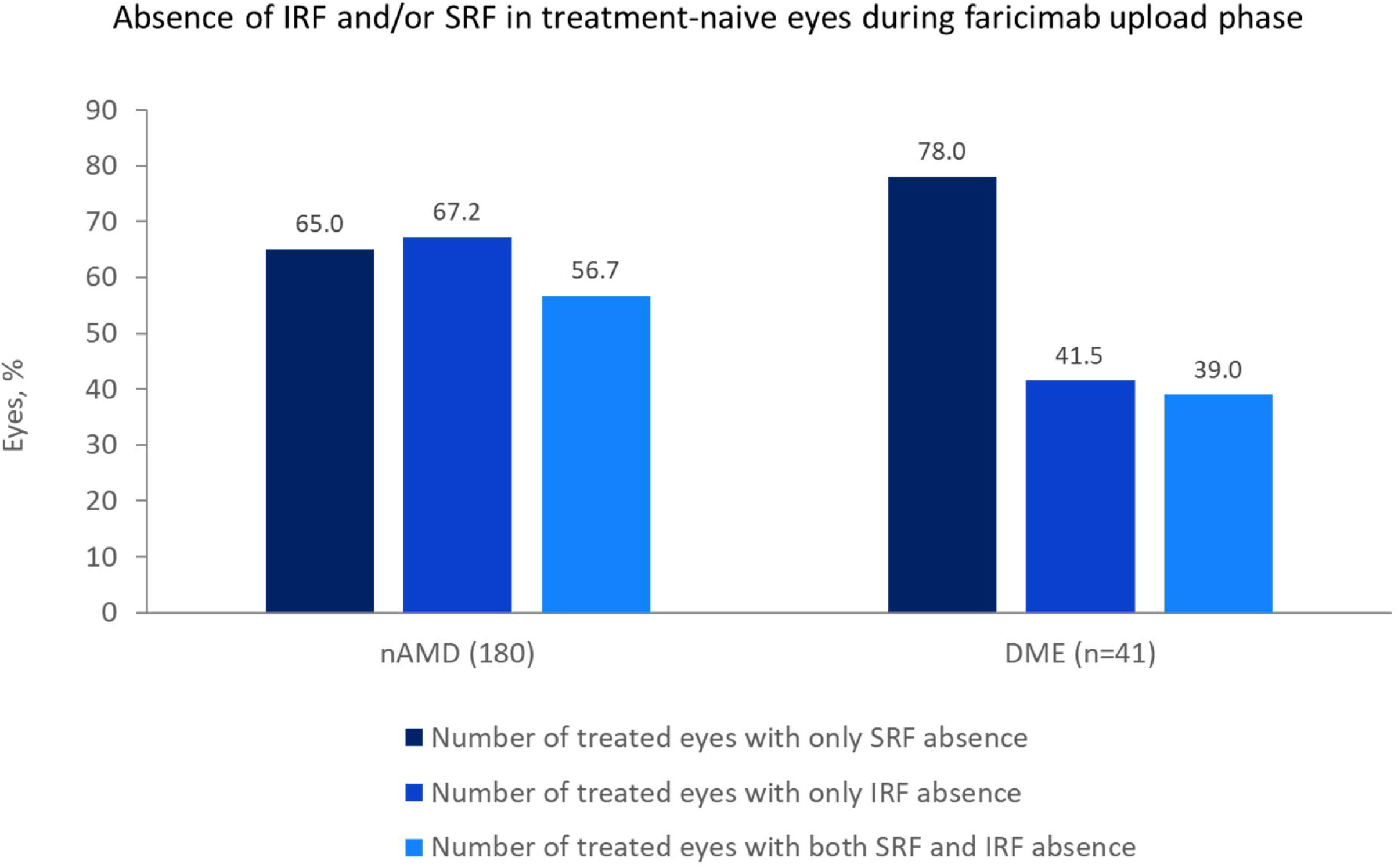
Absence of IRF and/or SRF in treatment-naive eyes during faricimab upload phase. N refers to the number of eyes. DME: diabetic macular edema; IRF: intraretinal fluid; nAMD: neovascular age-related macular degeneration; SRF: subretinal fluid

**Table 7:** Treatment-naive eyes with changes in decimal BDVA at site during faricimab upload phase.

|  | **nAMD**<br>(n=180) | **DME**<br>(n=41) |
| --- | --- | --- |
| **Changes in decimal BDVA, n (%)** |  |  |
| ≤0.1 | 62 (34.4) | 7 (17.1) |
| >0.1–0.2 | 33 (18.3) | 8 (19.5) |
| >0.2–0.3 | 27 (15.0) | 10 (24.4) |
| >0.3–0.4 | 32 (17.8) | 7 (17.1) |
| >0.4–0.5 | 6 (3.3) | 3 (7.3) |
| >0.5–0.6 | 7 (3.9) | 2 (4.9) |
| >0.6 | 13 (7.2) | 4 (9.8) |
| N refers to the number of eyes. BDVA: best-documented visual acuity; DME: diabetic macular edema; nAMD: neovascular age-related macular degeneration |  |  |

The main reasons for using faricimab as first-line therapy were to achieve a reduction in injection intervals (56.1 and 43.6% of nAMD and DME eyes, respectively) and best effectiveness outcome (42.9 vs 56.4). Overall, only 6.3% of 189 nAMD patients and 5.1% of 39 DME patients treated with faricimab in first-line discontinued faricimab treatment.

## DISCUSSION

This analysis provides valuable insights into routine care of ophthalmologic practice as well as first results for a large treatment-naive cohort treated with faricimab in Germany for nAMD as well as DME.

The first part of this retrospective analysis provides data on the predominant IVT regimens, drug upload strategies, and diagnostic imaging techniques commonly used in routine care of patients with nAMD and DME across ophthalmological practices and centers in Germany.

Analyzing IVT regimens applied at 24 German sites revealed that the majority of sites followed a treat-and-extend approach, regardless of the specific IVT administered. PRN was the second regimen of choice. This finding is consistent with previous registry analyses, reporting that treatment was mostly based on a treat-and-extend regimen (49% of patients) or PRN approach (46% of patients) (Wolfram, 2022). The choice of treatment regimens might be influenced by logistical factors such as patient availability, appointment scheduling constraints or practice processes and management. Altogether, the frequent use of T&E could indicate that sites prefer to determine treatment intervals individually for each patient and can be integrated well into the clinical practice process.

The initiation of an IVT injection therapy is commonly started with a so called upload, often refers to as 3-4 injections at intervals of 4 weeks, followed by the T&E/RPN regimen. With regard to upload strategies, the majority of sites did not distinguish between therapies when uploading and mostly applied three uploads in accordance with the summary of product characteristics (SmPC) and standard clinical practice. For faricimab treatment, sites preferred four IVT injections for the upload in adherence to the current SmPC (EMA, 2022). Still, some investigators reported to administer only one injection as an upload in their clinical practice. In this context, the research project did not evaluate the individual injection intervals of these patients after the upload of just one injection. Some might continue in a 4-week treatment rhythm, which technically equals the upload scheme.

Another important factor in IVT management is the use of imaging techniques. Current guidelines for the diagnosis and monitoring of nAMD and DME emphasize the central role of OCT in both diagnosis and treatment monitoring (Mowatt, 2014). The German Ophthalmological Society, the Retinological Society and the Professional Association of Ophthalmologists in Germany have pointed out in a statement on anti-VEGF therapy for nAMD that, in addition to an ophthalmological examination with best-corrected visual acuity, the diagnosis for IVT indication should also include a funduscopy with pupil dilation, OCT and, at least for initial indication, fluorescein angiography (G-BA, 2018). OCT was well established as standard procedure at therapy initiation in the clinical practice of all participating centers. Angiographic methods, such as FA, were still frequently used by two thirds of centers, supporting their continued relevance in clinical practice. However, OCT-A was used in only 25% of centers, likely due to limited availability or other restrictions. In this context other real-world studies demonstrated varying results (Modeste, 2024) where the application of OCT-A and FA may depend on individual clinical situation, device availability and insurance coverage which can also be applied to the situation in Germany.

The second part of this retrospective analysis provides data specifically for treatment-naive patients that initiate first-line therapy with faricimab up to upload completion as per physician’s definition.

The pivotal clinical trials for faricimab (YOSEMITE/RHINE (nAMD) and TENAYA/LUCERNE (DME)) demonstrated maintenance of visual improvements and robust anatomical outcomes with faricimab injection intervals of up to 16 weeks (Heier, 2022; Khanani, 2024; Quah, 2024; Wong, 2024). However, real-world data can display differences between clinical practice and controlled conditions of randomized controlled trials (RCTs) (Ciulla, 2021; Holz, 2015). In routine clinical settings, patients often receive fewer IVT injections and less frequent monitoring than conducted in RCTs. This discrepancy can be attributed to factors such as limited clinical capacities, logistical barriers, and patient compliance issues. Therefore, real-world evidence is crucial for understanding how these treatments perform in clinical practice. Importantly, a more heterogeneous patient population may be more challenging to treat and may lead to poorer outcomes due to significant ocular and systemic comorbidities.

Existing real-world studies on faricimab for nAMD (Kataoka, 2024; Khanani, 2023; Leung, 2023; Rush, 2023b) and DME (Kusuhara, 2023; Ohara, 2023; Rush, 2023a) supported its effectiveness in everyday clinical practice. However, these studies primarily focused on previously treated patients. So far, real-world data on faricimab therapy in treatment-naive nAMD and in particular DME patients are limited (Giancipoli, 2024; Kusuhara, 2023; Modeste, 2024; Nasimi, 2024; Penha, 2024; Quah, 2024) and have not yet been presented for a German cohort. Therefore, this is the first retrospective analysis placing special emphasis on previously IVT injection-naive nAMD and DME patients who started faricimab treatment in first-line since approval in Germany.

The ZEUS research project comprises a large patient cohort with 189 nAMD and 39 DME treatment-naive eyes, collected from 24 sites in Germany (including both practices and centers). Sites were selected which treated a considerable proportion of treatment-naive patients with faricimab as a first-line therapy. More than half of the sites treated ≥20 % of their treatment-naive nAMD patients with faricimab. In case of DME, 37% of sites treated at least 20% of treatment-naive patients with faricimab. In other studies, which did not focus on the recruitment of treatment-naive patients, fewer of such were included, as the majority of patients switched to faricimab after receiving another therapy (Heier, 2022; Khanani, 2023; Penha, 2024).

The collected baseline characteristics mainly align with those that have been published in pivotal as well as other real-world studies. Sex distribution with more females in the nAMD cohort aligns with a previous meta-analysis (Rudnicka, 2012), the pivotal trials TENAYA/LUCERNE (Khanani, 2024) and real-world study VOYAGER (Guymer, 2023) indicating that women have a higher risk of developing nAMD compared to men. Also higher numbers of male patients in the DME cohort are in line with numbers reported in YOSEMITE/RHINE (Wong, 2024) and VOYAGER (Guymer, 2023). In addition, males tend to show a higher prevalence of severe diabetic retinopathy, likely due to poorer metabolic control and the resulting diabetic maculopathy (Mathur, 2017). Additionally, the age distribution matched with previous reports, with nAMD patients being older than those with DME (Guymer, 2023; Penha, 2024; Quah, 2024). The higher prevalence of type II diabetes patients (74.4%) in DME is consistent with findings in other studies (type II: 84.5%) (Quah, 2024). The high number of DRIL (48%) at baseline in treatment-naïve DME eyes might suggests that these patients were in a more advanced stage of the disease than expected in a cohort of untreated patients. Studies have shown that the presence of DRIL correlates with worse visual outcomes (Das, 2018), indicating that treatment-naive eyes may already exhibit significant retinal damage at the time of diagnosis, which might be an additional factor to choose a potent dual mechanism of action. In contrast to clinical trials (TENAYA/LUCERNE: 3.5%) (Khanani, 2024), almost one in three patients (28.6%) had an unknown classification of CNV, that may reflect differences in diagnostic criteria or imaging limitations. Low numbers of nAMD patients with RAP might result from infrequent performance of fluorescein angiography.

The occurrence of IRF and/or SRF is a typical feature of nAMD and DME. The proportion of patients with IRF (67.8%) or SRF (65.3%) at baseline in the nAMD population was comparable to the VOYAGER study (IRF/SRF: 62.7%/69.8%) (Guymer, 2023). As expected, more eyes had only IRF (77.1%) in DME compared to nAMD eyes. The ratio of IRF to SRF is comparable to data reported in the VOYAGER study (IRF/SRF: 98.2%/29.8%) (Guymer, 2023) and real-world studies (e.g., IRF/SRF: 100 %/30.3 %) (Quah, 2024). Of note, CRT not always displayed the localization of greatest fluid volume. Although CRT is used as a standardized parameter, it seems not always to be representative of highest fluid accumulation. It is important to consider, that patients with a low CRT value can still have significant fluid accumulation in other regions of the retina. This may explain why some patients experience a decline in visual acuity or disease progression despite a seemingly stable CRT.

For the longitudinal analysis of treatment-naive faricimab patients CRT reduction, absence of fluid and and changes in visual acuity were assessed after completion of upload per physician’s definition. The majority of both, nAMD and DME patients received 4 upload IVT injections as per label. However, as also observed in the analysis of the general clinical practice, a certain proportion (22.9% for nAMD and 16.7% for DME) of the patients received an upload of only one IVT injection.

During the faricimab upload phase, one-third of patients with nAMD achieved a reduction in CRT of over 60%, exceeding the findings from the FARETINA study, reporting a mean CRT reduction of 15% after the fourth IVT injection in treatment-naive patients (Lim, 2024). Patients with DME experienced a less pronounced CRT reduction, but improvement was higher compared to the FARETINA trial demonstrating a 9% CRT reduction after the fourth IVT injection (Sheth, 2024). However, it needs to be considered that due to the aggregated data collection, no baseline CRT reference value was determined which complicates the clear interpretation of the described percentual reduction.

More than 65% of nAMD patients achieved absence of IRF and SRF. These numbers were lower compared to pivotal trials showing IRF and/or SRF absence of 88% (for IRF, SRF) and 77 % (for IRF and SRF) after 12 weeks of faricimab treatment (TENAYA/LUCERNE) (Khanani, 2024). However, it needs to be considered that some patients received less than four upload injections. Thus, shorter upload intervals and resulting treatment durations of less than 12 weeks might explain lower numbers of patients achieving IRF/SRF absence so far. The variability in injection frequency among centers may suggest a need for standardized real-world treatment protocols.

Besides CRT reduction and absence of retinal fluids, visual acuity is an important outcome parameter since a positive effect on vision is the ultimate treatment goal directly affecting patients’ daily life. Despite heterogeneous upload intervals, visual acuity was improved in most nAMD and DME treatment-naive eyes during faricimab upload phase. Physicians documented the decimal visual acuity, that technically can be converted into an ETDRS score as that a step of 0.1 −log10(VAdec) corresponds to 1 line in ETDRS score (5 letters). However, conversion is non-linear and depends on the baseline BDVA that was very broadly distributed. An improvement of 0.2-0.3 in decimal visual acuity corresponds to an improvement of 8 - 12 in the ETDRS for patients with a baseline decimal visus of 0.4, which approximately corresponds to the median baseline visus of the nAMD patients. It also needs to be considered that the majority of nAMD eyes (56.1 %) initiated treatment with relatively good visual acuity of >0.4 (equaling >65 ETDRS letters) which could explain why 34% of the eyes showed smaller improvements of ≤0.1 in decimal visual acuity during the upload phase due to a ceiling effect.

DME patients received more loading doses, suggesting that DME is more challenging to treat, requiring more injections to achieve major improvements (Giancipoli, 2024). With its complex metabolic and inflammatory processes, the underlying type II diabetes could explain the challenge (Sakini, 2024). Physiologically, the continuously activated Ang-1/Tie-2 signalling cascade functions as a “molecular brake” against inflammation (G-BA, 2022a; Heier, 2021). Under certain conditions, Ang-2 levels are increased and Ang-2 acts as competitive antagonist of Ang-1 by binding to the Tie2 receptor, promoting proinflammatory responses (Chaudhary, 2024; Heier, 2021). Thus, the anti-inflammatory properties of faricimab in nAMD and DME through Ang-2 inhibition might favor it as a first-line treatment option.

Limitations of this analysis include its retrospective and descriptive nature, limited racial and ethnic diversity, and reliance on available case notes and OCT data. We are aware that data were obtained from a limited number of 24 centers, that were selected with the aim to include a considerable number of treatment-naive patients. In addition, the decision which treatment-naive patients to include was with the investigators (within specified inclusion and exclusion criteria). Therefore, results might not fully reflect the overall treatment situation in Germany. Reliance on aggregated data without patient-level follow-up did not allow statistical analysis to determine any correlation or significance. A limitation for analysis of treatment regimens and upload injections in general is the lack of differentiation based on the specific indications nAMD versus DME, which could provide further insights into treatment regimen selection and upload modalities. Further limitations to consider include that follow-up timelines were not available and patients may have been on faricimab therapy for varying amounts of follow-up. Predefined categories were specified for the evaluation of the CRT and visual acuity results, and IRF/SRF and PED was evaluated as presence/absence, while reduction was not analysed. In addition, heterogenous upload regimes have to be considered when interpreting longitudinal data. Longitudinal outcomes presented here are reported from patients with diverse injection patterns (1-4 injections) which limits interpretability since not all patients were measured at the same time point and received the same number of faricimab injections. Therefore, the analysis timepoint for longitudinal endpoints may be considered as “during upload phase”. While the findings align with controlled clinical trials, the absence of standardized follow-up intervals in real-world practice may have influenced the reported outcomes. Adverse events were not recorded in this research project. However, the low number of patients discontinuing treatment indicates that faricimab was well tolerated as seen in pivotal trials. To minimize bias, all sites underwent standardized training and utilized standardized documentation for completing of eCRFs. To reduce the potential bias by retrospective data use, in- and exclusion criteria were defined to include patients representing the site’s general population regarding patient disposition and IVT regimen, and to minimize missing data. Besides all limitations, the current data analysis provides valuable insights into clinical practice patterns of IVT application based on a large cohort of patients with nAMD and DME.

## CONCLUSION

This study provides the first real-world evidence on faricimab in a large cohort of treatment-naive nAMD and DME patients in Germany. T&E treatment regimens were frequently used regardless of intravitreal therapy, which mirrors the preference of individual approaches based on patients’ needs. The data support the use of faricimab as a first-line therapy resulting in drying and acuity gain during upload; future studies should explore long-term outcomes and optimal retreatment strategies.

## Data Availability

All data produced in the present study are available upon reasonable request to the authors.

## Acknowledgements

The authors would like to thank all investigators study centers involved in this research project: Jan Alder (Dr. Dierse & Partner), Albert Augustin (Privatärztliche Gemeinschaftspraxis Dr. Scholl & Prof. Augustin), Julia Clauß (Augenzentrum am Johannisplatz), Roxana Fulga (BeyondEye GmbH), Andreas Georg Gossen (Evang. Kliniken Essen Mitte), Janek Häntzschel (Augenzentrum Pirna MVZ), Frederike Hochhaus (Augenzentrum Erding GbR), Waldemar Jendritza (Gemeinschaftspraxis Dr. Bettina und Waldemar Jendritza), Hakan Kaymak (MVZ Breyer, Kaymak & Klabe, Oberkassel, Internationale Innovative Ophthalmochirurgie GbR), Christoph Kern (Augenärzte Oberland PD Dr. Dinslage & PD Dr. Kern GbR), Marcus Kernt (Augenarztpraxis Prof. Dr. med. habil. Marcus Kernt), Adnan Kilani (Universitäts-Augenklinik Ulm, Prof. Armin Wolf), Jonathan Krauter (Artemis Augenarzt-Praxis Babenhausen), Victoria Linse (Augenarztpraxis Dr. Herbrig), Laura Lux (Augenzentrum Cottbus BAG), Melika Mirdamadi (Augenarztpraxis am Elsterplatz), Anna-Katharina Müller (Augenzentrum Cottbus BAG), Gudrun Papadopoulos (Augenzentrum Hochrhein), Lisa Pelz (Praxis Dr. Bettina Kahle), Thilo Schimitzek (Augenklinik Kempten), Ralf Schmitt (Augenzentrum Saarbrücken im Medizeum), Wolfgang Schrader (Augenzentrum Würzburg MVZ GmbH), Ricarda G. Schumann (Augenzentrum München Schwabing), Finn Theine (AZSH Neumünster), Ulrich Thelen (Augenärzte Klosterstraße). Medical writing assistance was provided by med:unit GmbH, Germany, and was funded by Roche Pharma AG. The authors had full editorial control and gave their final approval.

